# Different Combinations of the Metabolic Syndrome Criteria and Cardiovascular Disease

**DOI:** 10.1101/2023.10.11.23296877

**Authors:** Lars Lind

## Abstract

**Background:** The metabolic syndrome (MetS) has previously been linked to incident cardiovascular disease (CVD). It is however not known if certain combinations of MetS criteria show a higher risk than others.

**Methods:** We used data from UK biobank in which 388,800 individuals had data on MetS using the five harmonized NCEP criteria. The cohort was followed for a median of 12.6 years. A composite CVD outcome was used (myocardial infarction, ischemic stroke or heart failure).

**Results:** The risk of incident CVD (n= 22,572) increased in a fairly linear fashion with increasing number of MetS criteria. In the groups showing three MetS criteria, thus fulfilling the definition of MetS, the highest risk was seen in those with the combination of the glucose + waist circumference + HDL criteria (HR 3.90, 95%CI 3.17-4.80). In the group with four criteria, the highest HRs were seen in the groups not including triglyceride or the blood pressure criteria (HR 3.96, 95%CI 3.57-4.59 and HR 3.79, 95%CI 3.13-4.59, respectively). The risk of CVD in those with all five criteria was in the same order (HR 3.65, 95%CI 3.33-4.01). Mendelian randomization indicated a causal role of MetS for coronary heart disease (CHD) and heart failure, but not ischemic stroke, while use of polygenetic risk scores for CHD and ischemic stroke were related to MetS criteria in a similar fashion as observational data.

**Conclusion:** Certain combinations of risk factors in individuals with the metabolic syndrome criteria showed a higher risk of future CVD than others.

## INTRODUCTION

The metabolic syndrome (MetS) was in modern times described in 1988 ^1, 2^. Although it is widely used in the clinic to denote the abdominal obese subject with cardio-metabolic disturbances, it has been questioned as a pathophysiological entity ^3^, since MetS does not show any increased risk of cardiovascular disease (CVD) as compared with the sum of its components ^4^. Another critique is that given its 5 different criteria, a subject with MetS could have 15 different combinations of these criteria and it is likely that certain combinations are more deleterious than others.

Many previous studies have shown MetS, and the number of MetS components, to be related to future CVD and/or mortality ^3, 5–17^. It has also been shown that the impact of MetS as a risk factor declines by ageing, although it is still a significant risk factor at the age of 82 years ^18^. However, since most studies lack the power to investigate if different combinations of the MetS criteria have different impact on CVD risk, this is a matter that needs to be evaluated.

The present study was therefore undertaken with the primary aim to investigate if different combinations of the MetS criteria show a higher risk than others in terms of future CVD. For this purpose, we used the UK biobank, a study with a large sample size and a long follow-up period that has sufficient power to be used for this aim. A secondary aim was to evaluate if different combinations of the MetS criteria are more deleterious than others regarding the risk of future specific major CVDs, like myocardial infarction, stroke, or heart failure. A third aim was to perform a two-sample Mendelian randomization study to evaluate if the relationships between MetS and the three major CVDs were causal. For this aim, we also used derived polygenetic risk scores (PRS) ^19^. The primary hypothesis tested was that different combinations of MetS criteria show a higher risk than others regarding incident CVD.

## MATERIAL AND METHODS

### Sample

UK Biobank is a large, multi-center, prospective cohort study conducted across the UK (https://www.ukbiobank.ac.uk). In 2006–10, over 500,000 individuals aged 40–69 years underwent physical measurements, and blood samples were biobanked for later analysis of genes and biomarkers. After excluding individuals with missing data on variables needed to define MetS, 388,800 remained eligible for analysis.

The study was approved by the UK North West Multi-Centre Research Ethics Committee and the Swedish Ethical Review Authority. All participants provided written informed consent.

### Clinical and biochemical data

Serum glucose, HDL-cholesterol, and triglycerides were measured by a Beckman coulter AU5800, by standard methods. Since samples were taken with different times of fasting, glucose levels were adjusted down by 1.5 mmol/l if reported fasting time was 0h, - 3.0 mmol/l if fasting was 1h, -1.0 mmol/l if fasting was 2h, -0.3 mmol/l if fasting was 3h and no correction if fasting time was >3h. For triglycerides, the levels were adjusted down by 0.1 mmol/l if reported fasting time was 1h, and the reductions were -0.2, -0.4, -0.6, -0.65, -0.4 and -0.1 mmol/l for times 2 to 7h, respectively. These adjustments are based on a literature search, as well as own experience of the response to a mixed meal. Blood pressure was measured twice in the sitting position with the automated Omron device.

### Confounders

Ethnicity was categorized into four groups; White, Black, Asian, and other. Townsend social deprivation index and income were used as markers of socioeconomic status. Use of statins was regarded as a marker of high LDL-cholesterol (not being a criterion of MetS).

### The metabolic syndrome

The harmonized National Cholesterol Education Program (NCEP)-criteria for MetS was used to define the 5 components of the syndrome and prevalent MetS (binary) ^20^. Three of the following five criteria should be fulfilled: Blood pressure ≥ 130/85 mmHg or antihypertensive treatment, serum glucose ≥ 6.1 mmol/l or antidiabetic treatment, serum triglycerides ≥ 1.7 mmol/l, waist circumference > 102 cm in men and > 88 cm in women, HDL-cholesterol < 1.0 mmol/l in men and < 1.3 in women.

### Outcomes

First fatal or non-fatal myocardial infarction (ICD-10 code I21), ischemic stroke (I63), or heart failure (I50) were recorded from death certificates and hospital records.

### Statistics

Cox proportional hazard analysis was used to evaluate if MetS, number of MetS criteria, or MetS criteria combinations were related to incident CVD (combined endpoint of myocardial infarction, ischemic stroke, or heart failure) in the primary analysis. Confounders were age, sex, ethnicity, Townsend deprivation index, smoking (never, previous, current), and statin use.

Prevalent CVD at baseline was deleted before analyses.

The proportional hazard assumption was checked by visual inspection of Kaplan-Maier curves.

As secondary outcomes, we evaluated in a similar fashion the three CVDs myocardial infarction, ischemic stroke, or heart failure one by one.

Two-sample Mendelian randomization studies were performed using already published genome-wide association studies (GWAS) for the three major CVDs (CARDIOGRAMplusC4D for coronary heart disease ^21^, HERMES for heart failure ^22^, METASTROKE for ischemic stroke ^11^, and a GWAS of MetS performed in the UK biobank (the exposure) ^23^. As instruments for the exposure, only independent genetic loci with p<5 x 10^-8^ were used. Independency of the SNPs was evaluated by the clump command in the MRbase package in R. Thereafter, the causal estimate was calculated by both inverse-variance weighted meta-analysis (IVW) and MR Egger, as well as the weighted-median method. We regarded a significant IVW estimate to be causal if also the weighted-median estimate was significant, and the MR Egger estimate to be in the same order as the other two.

Weale and co-workers have constructed polygenetic risk scores (PRS) for a number of traits, including coronary heart disease (CHD) and ischemic stroke, for the participants in UK biobank ^19^. The PRSs for these two CVD outcomes were calculated using published GWAS (called standard PRS). The PRSs were calculated using meta-analyses using a Bayesian approach to estimate non-zero weights in ∼6M SNPs spread throughout the genome (see more information in Supplementary Tables 1-4 in the publication). The individual PRS values were calculated as the sum of the effect size multiplied by allele dosage. Thereafter, the PRSs were centered and standardized in order to achieve PRS values that were comparable between traits. When the different groups with MetS combinations were related to the PRSs for CHD and ischemic stroke, linear regression models were used with the same confounders as used above when evaluating incident outcomes.

STATA16.1 was used for the analyses (Stata inc, College Station, TX, USA).

## RESULTS

Basic characteristics in the sample are shown in Table 1.

**Table 1.**
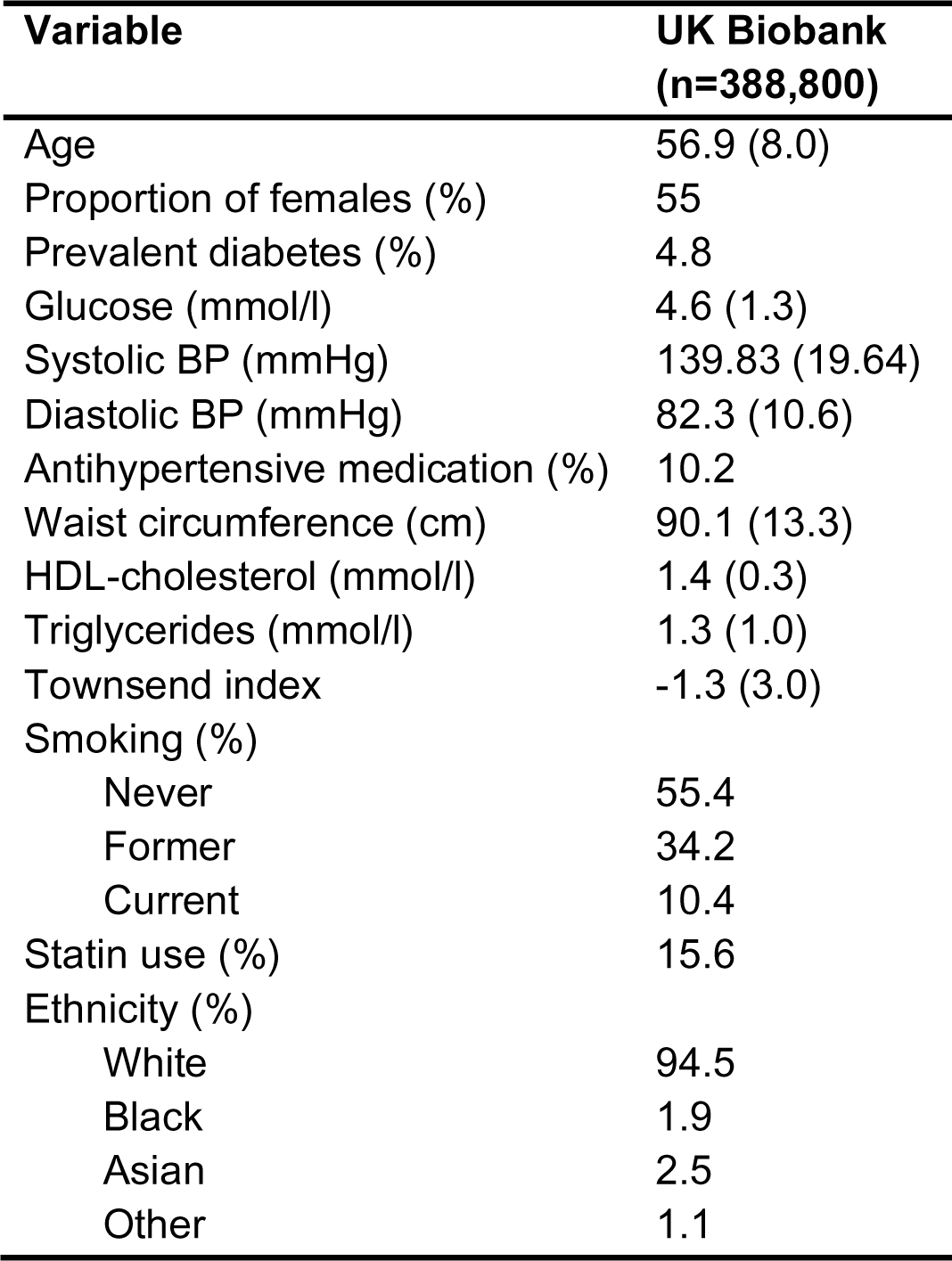
Basic characteristics in the sample with complete data on metabolic syndrome components and after exclusion of individuals with prevalent cardiovascular diseases at the baseline investigation (n=388,801). Means and SD are given or proportions.

| **Variable** | **UK Biobank**<br>**(n=388,800)** |
| --- | --- |
| Age | 56.9 (8.0) |
| Proportion of females (%) | 55 |
| Prevalent diabetes (%) | 4.8 |
| Glucose (mmol/l) | 4.6 (1.3) |
| Systolic BP (mmHg) | 139.83 (19.64) |
| Diastolic BP (mmHg) | 82.3 (10.6) |
| Antihypertensive medication (%) | 10.2 |
| Waist circumference (cm) | 90.1 (13.3) |
| HDL-cholesterol (mmol/l) | 1.4 (0.3) |
| Triglycerides (mmol/l) | 1.3 (1.0) |
| Townsend index | -1.3 (3.0) |
| Smoking (%) |  |
| Never | 55.4 |
| Former | 34.2 |
| Current | 10.4 |
| Statin use (%) | 15.6 |
| Ethnicity (%) |  |
| White | 94.5 |
| Black | 1.9 |
| Asian | 2.5 |
| Other | 1.1 |

### Incidence rate for CVD

During a median follow-up of 12.6 years (max 14.9), 22,572 individuals experienced a first CVD event, resulting in an incidence rate of 4.8 per 1000 person years at risk (PYAR). The corresponding number of incident events for myocardial infarction, heart failure, and ischemic stroke were 9,315, 6,425, and 12,385, respectively.

### MetS and CVD

20.6% of the sample showed MetS. The incidence of the combined endpoint CVD was roughly twice in individuals with MetS compared to those without (Table 2). Following adjustment for age, sex, and other confounders the HR was 1.59 (95% 1.55-1.64, Table 3). When investigating the three CVD diseases separately, the HRs for MI and HF were similar (1.64 and 1.63, respectively), while the HR for ischemic stroke was lower (1.45, see Table 3 for details).

**Table 2.** Number of individuals, number (and %) of incident CVD events (composite endpoint) during the follow-up period when the sample was divided into those with or without the metabolic syndrome (MetS), or the number of MetS criteria, or when the sample was subdivided into the different combinations of the five criteria of MetS. GLU=glucose criteria, BP=blood pressure criteria, TG=triglyceride criteria, HDL= HDL-cholesterol criteria, WC= waist circumference criteria.

|  | **No. of**<br>**individuals** | **No. of**<br>**incident**<br>**CVD** | **% with**<br>**incident**<br>**CVD** |
| --- | --- | --- | --- |
| **MetS (binary)** |  |  |  |
| 0 | 308,889 | 15,021 | 4.86% |
| 1 | 79,912 | 7,552 | 9.45% |
| **No. of MetS criteria** |  |  |  |
| 0 | 64,570 | 1,317 | 2.04% |
| 1 | 147,018 | 7,059 | 4.80% |
| 2 | 97,301 | 6,645 | 6.83% |
| 3 | 53,791 | 4,509 | 8.38% |
| 4 | 22,087 | 2,417 | 10.94% |
| 5 | 4,034 | 626 | 15.52% |
| **MetS criteria combinations** |  |  |  |
| No criterion | 63,198 | 1,284 | 2.03% |
| Only GLU | 2,378 | 91 | 3.83% |
| Only BP | 113,027 | 5,708 | 5.05% |
| Only TG | 7,947 | 274 | 3.45% |
| Only HDL | 2,356 | 120 | 5.09% |
| Only WC | 8,921 | 337 | 3.78% |
| GLU+BP | 10,797 | 797 | 7.38% |
| GLU+TG | 517 | 45 | 8.70% |
| GLU+HDL | 648 | 44 | 6.79% |
| GLU+WC | 992 | 92 | 9.27% |
| BP+WC | 36,249 | 2,584 | 7.13% |
| BP+HDL | 12,089 | 809 | 6.69% |
| BP+TG | 26,898 | 1,767 | 6.57% |
| WC+TG | 2,782 | 119 | 4.28% |
| WC+HD | 3,698 | 150 | 4.06% |
| TG+HDL | 7,466 | 372 | 4.98% |
| GLU+BP+WC | 6,547 | 803 | 12.27% |
| GLU+BP+TG | 3,162 | 297 | 9.39% |
| GLU+BP+HDL | 1,691 | 189 | 11.18% |
| GLU+WC+TG | 424 | 45 | 10.61% |
| GLU+WC+HDL | 748 | 96 | 12.83% |
| GLU+TG+HDL | 445 | 43 | 9.66% |
| BP+WC+TG | 16,937 | 1,330 | 7.85% |
| BP+WC+HDL | 11,154 | 842 | 7.55% |
| BP+TG+HDL | 11,587 | 887 | 7.66% |
| Not GLU | 13,960 | 1,244 | 8.91% |
| Not HDL | 3,632 | 467 | 12.86% |
| Not TG | 3,479 | 516 | 14.83% |
| Not BP | 884 | 116 | 13.12% |
| Not WC | 1,853 | 213 | 11.49% |
| All five criteria | 5,108 | 721 | 14.12% |

**Table 3.** Relationships between having the metabolic syndrome (MetS), or having a certain number of MetS criteria, and of incident CVD events (composite endpoint, primary analysis) during the follow-up period. Those with MetS were compared to those without, while the different number of MetS criteria were all compared to individuals without any criteria. Also other endpoints are given in the same fashion.

|  |  | **HR** | **95%CI** | **SE** | **Wald.** | **P-value** |
| --- | --- | --- | --- | --- | --- | --- |
| **CVD (composite endpoint)** |  |  |  |  |  |  |
| **MetS binary** |  | **1.59** | **1.55-1.64** | **0.02** | **32.17** | **<0.0001** |
| MetS no. of criteria | 1 | 1.60 | 1.50-1.69 | 0.05 | 15.43 | <0.0001 |
|  | 2 | 2.06 | 1.94-2.18 | 0.06 | 23.61 | <0.0001 |
|  | 3 | 2.44 | 2.29-2.59 | 0.08 | 28.00 | <0.0001 |
|  | 4 | 3.12 | 2.91-3.34 | 0.11 | 32.64 | <0.0001 |
|  | 5 | 3.65 | 3.33-4.01 | 0.19 | 27.04 | <0.0001 |
| **Myocardial infarction** |  |  |  |  |  |  |
| **MetS binary** |  | **1.65** | **1.58-1.72** | **0.04** | **22.25** | **<0.0001** |
| MetS no. of criteria | 1 | 1.82 | 1.65-2.00 | 0.09 | 12.27 | <0.0001 |
|  | 2 | 2.31 | 2.10-2.55 | 0.11 | 17.08 | <0.0001 |
|  | 3 | 2.84 | 2.57-3.14 | 0.14 | 20.53 | <0.0001 |
|  | 4 | 3.56 | 3.20-3.97 | 0.20 | 22.86 | <0.0001 |
|  | 5 | 4.12 | 3.56-4.77 | 0.34 | 18.86 | <0.0001 |
| **Heart failure** |  |  |  |  |  |  |
| **MetS binary** |  | **1.63** | **1.57-1.69** | **0.03** | **25.59** | **<0.0001** |
| MetS no. of criteria | 1 | 1.35 | 1.24-1.46 | 0.06 | 7.18 | <0.0001 |
|  | 2 | 1.78 | 1.64-1.93 | 0.07 | 13.81 | <0.0001 |
|  | 3 | 2.13 | 1.96-2.32 | 0.09 | 17.57 | <0.0001 |
|  | 4 | 2.81 | 2.57-3.08 | 0.13 | 22.31 | <0.0001 |
|  | 5 | 3.48 | 3.10-3.91 | 0.22 | 20.63 | <0.0001 |
| **Ischemic stroke** |  |  |  |  |  |  |
| **MetS binary** |  | **1.45** | **1.38-1.53** | **0.04** | **13.68** | **<0.0001** |
| MetS no. of criteria | 1 | 1.48 | 1.33-1.65 | 0.08 | 7.23 | <0.0001 |
|  | 2 | 1.72 | 1.54-1.91 | 0.09 | 9.82 | <0.0001 |
|  | 3 | 1.97 | 1.76-2.20 | 0.11 | 11.69 | <0.0001 |
|  | 4 | 2.50 | 2.20-2.83 | 0.16 | 14.36 | <0.0001 |
|  | 5 | 3.26 | 2.76-3.85 | 0.30 | 13.95 | <0.0001 |

### Number of MetS criteria and CVD

The proportion of incident CVD cases increased by a factor of 7.5 when the group with no MetS criterion was compared with those with all five criteria. As could be seen in Figure 1 and Table 3, the HR risk of CVD increased in a fairly linear fashion with the number of MetS criteria. Following adjustment for age, sex, and other confounders the HR was 3.65 (95% 3.33-4.01, Table 3) in the group with all five criteria compared to the group with no criterion. When investigating the three CVD diseases separately, HR in the group with 5 criteria was 4.12 regarding MI, but not as high for the outcomes stroke and heart failure (3.26 and 3.48) when compared to the group with no criterion.

**Figure 1.**
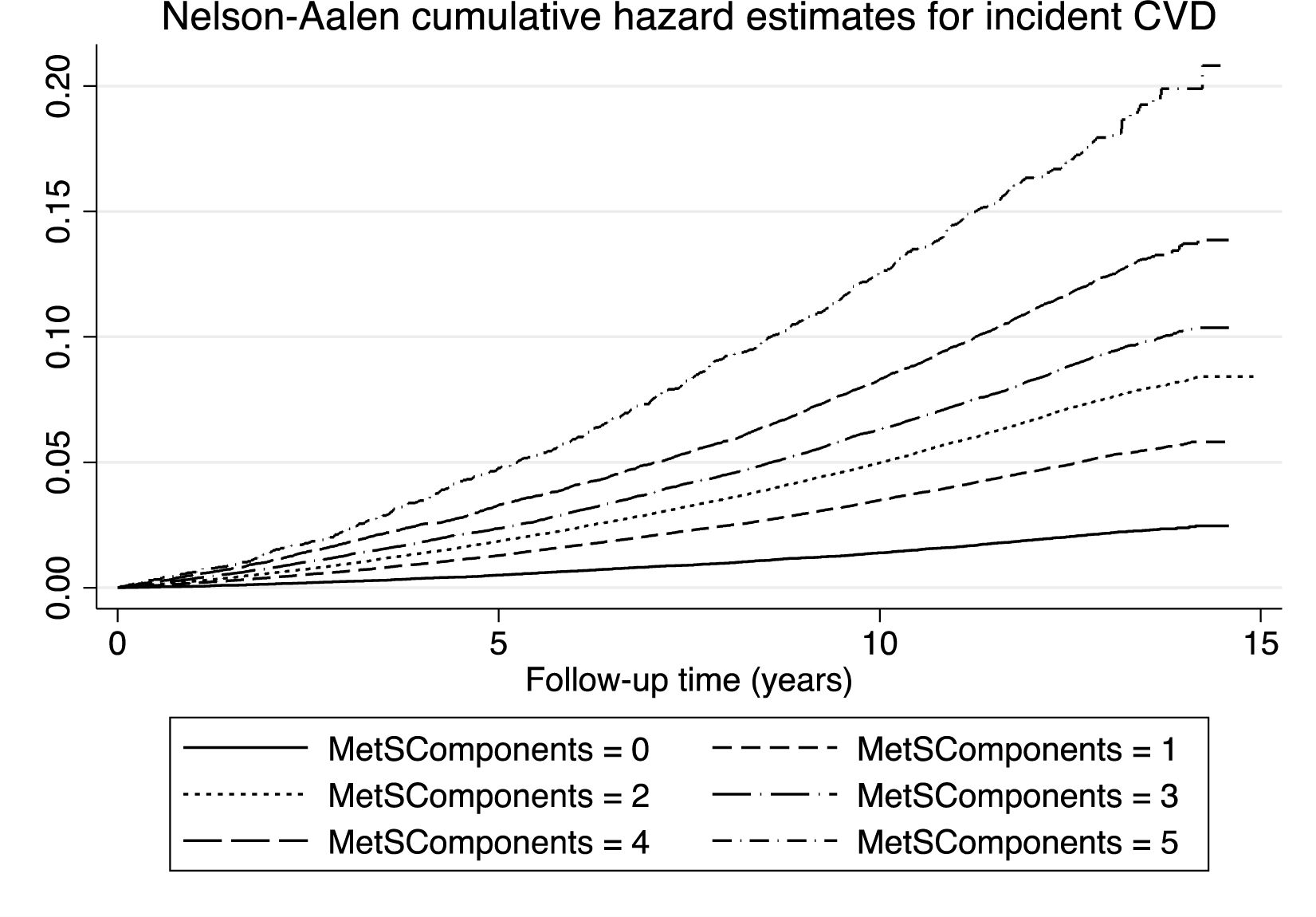
Cumulative hazard estimates for groups with different number of metabolic syndrome criteria (components) regarding the composite CVD endpoint during the follow-up period. P<0.0001 for overall difference between groups.

### Combinations of MetS criteria and CVD

As could be seen in Figure 2, the 30 possible different combinations of criteria were compared to subjects without any MetS criteria. It was found that the spread in HRs between the 30 groups of MetS criteria combinations were large, also within groups with a similar number of criteria. Amongst subjects with one criterion, the group with the blood pressure (BP) criterion showed the highest HR when compared to the group without any criterion, and the group with the triglyceride (TG) criterion showed the lowest HR. All these 5 groups did however show an increased risk of incident CVD (see Figure 3 and Suppl Table 1). In general, the groups with 2 criteria showed higher HRs than those with one criterion only. Within this category, the highest HR was seen for the combination of glucose (GLU) + waist circumference (WC) (2.71) and the lowest HR in those with WC+TG (1.51). In the groups showing 3 MetS criteria, thus fulfilling the definition of MetS, the highest HR was seen in those with GLU+WC+HDL (HR 3.90), while the lowest HRs were seen for three combinations all including TG (HRs 2.11 to 2.21). In the group with 4 components, the highest HR was seen in the groups not including TG or BP (3.96 and 3.79, respectively), while the lowest HR was seen in the group not including WC. It is also of interest to see that the HR for those with all 5 criteria was in the same order as in those with 4 criteria not including TG or BP, as well as the group with GLU+WC+HDL, all with overlapping confidence intervals.

**Figure 2.**
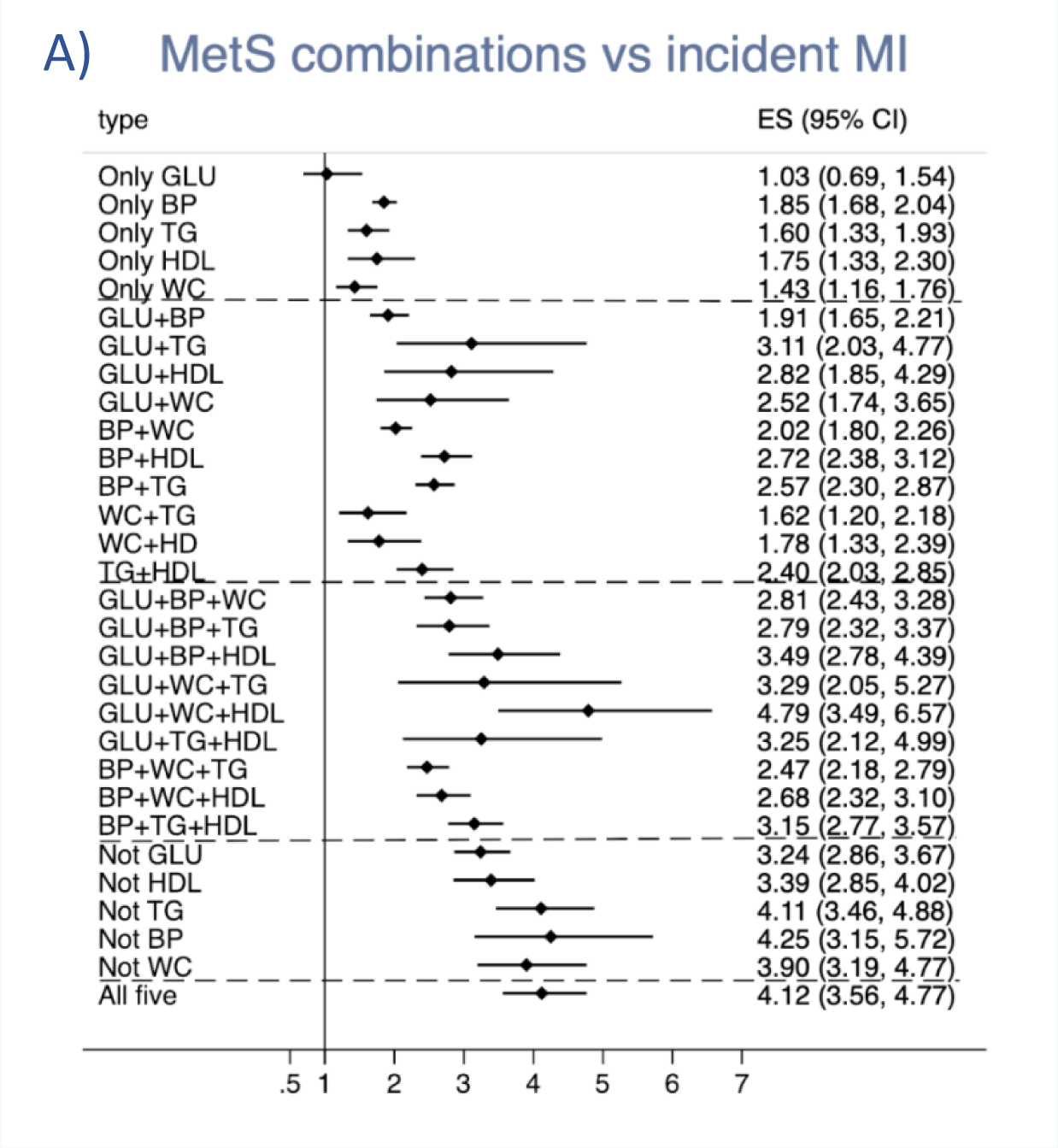
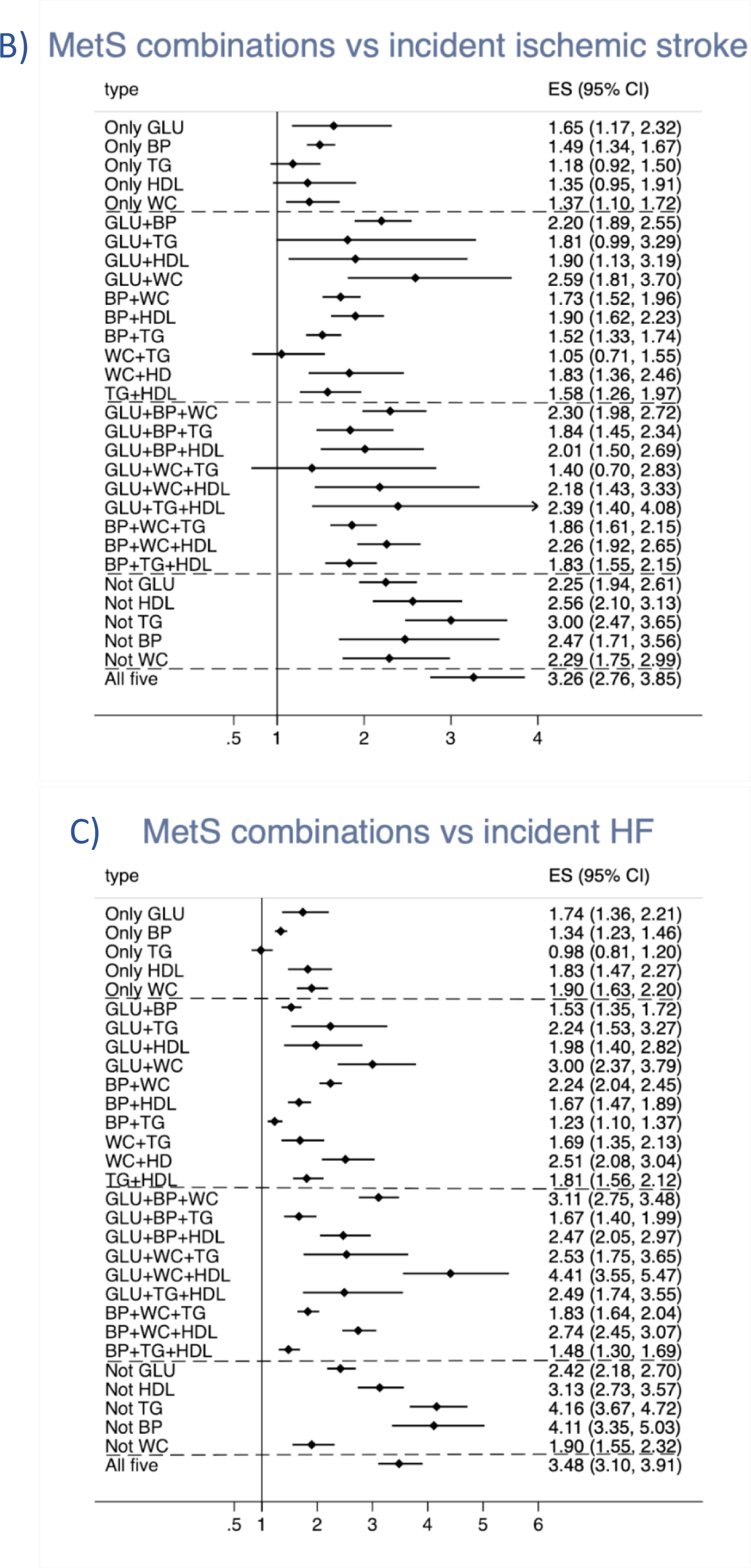
Relationships between different combinations of the five metabolic syndrome (MetS) criteria and (A) incident myocardial infarction, (B) ischemic stroke, and (C) heart failure. The estimate (ES) is the hazard ratio for comparison vs the group of individuals without any MetS component. GLU=glucose criteria, BP=blood pressure criteria, TG=triglyceride criteria, HDL= HDL-cholesterol criteria, WC= waist circumference criteria. The dashed lines denote the divisions between 1, 2, 3, 4, and 5 criteria.

**Figure 3.**
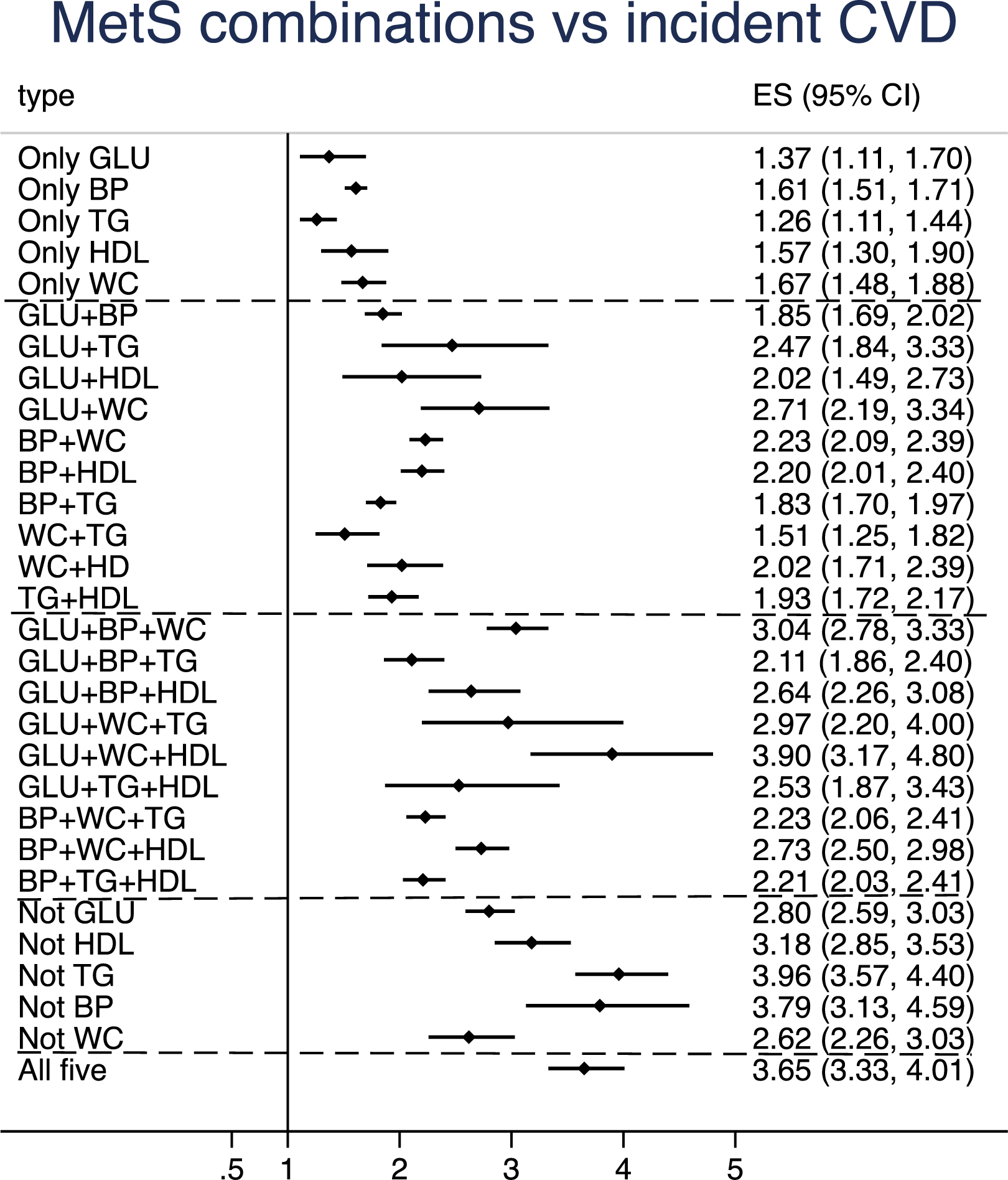
Relationships between different combinations of the five metabolic syndrome (MetS) criteria and incident cardiovascular disease (CVD) (combined endpoint): The estimate (ES) is the hazard ratio for comparison vs the group of individuals without any MetS component. GLU=glucose criteria, BP=blood pressure criteria, TG=triglyceride criteria, HDL= HDL-cholesterol criteria, WC= waist circumference criteria. The dashed lines denote the division between 1, 2, 3, 4, and 5 criteria.

When the pattern of HRs for the 30 MetS combination groups was compared between the three different CVDs, the pattern for MI was the CVD most similar with the pattern for the combined endpoint CVD with high HRs for those with all 5 criteria, those with 4 criteria not including TG or BP and in the group with GLU+WC+HDL. Those 3 groups were also found to have the highest HRs for the endpoint MI, but in that case also those with 4 criteria not including WC showed a similar HR. Regarding ischemic stroke, the groups with all 5 criteria and those with 4 criteria not including TG showed the highest HRs, while the GLU+WC+HDL group was not as prominent as for MI and HF.

### Mendelian randomization (MR)

76 independent SNPs from the GWAS of MetS were used as instrumental variables for the exposure in the MR (see reference 22 for details on these SNPs). As could be seen in Table 4, all three tests showed a causal effect of MetS on coronary heart disease (IVW estimate 1.25, 95%CI 1.20-1.29, p=1.0*10^-10^) and heart failure (IVW estimate 1.13, 95%CI 1.10-1.17, p=8.9*10^-17^), but were not significant regarding ischemic stroke.

**Table 4.**
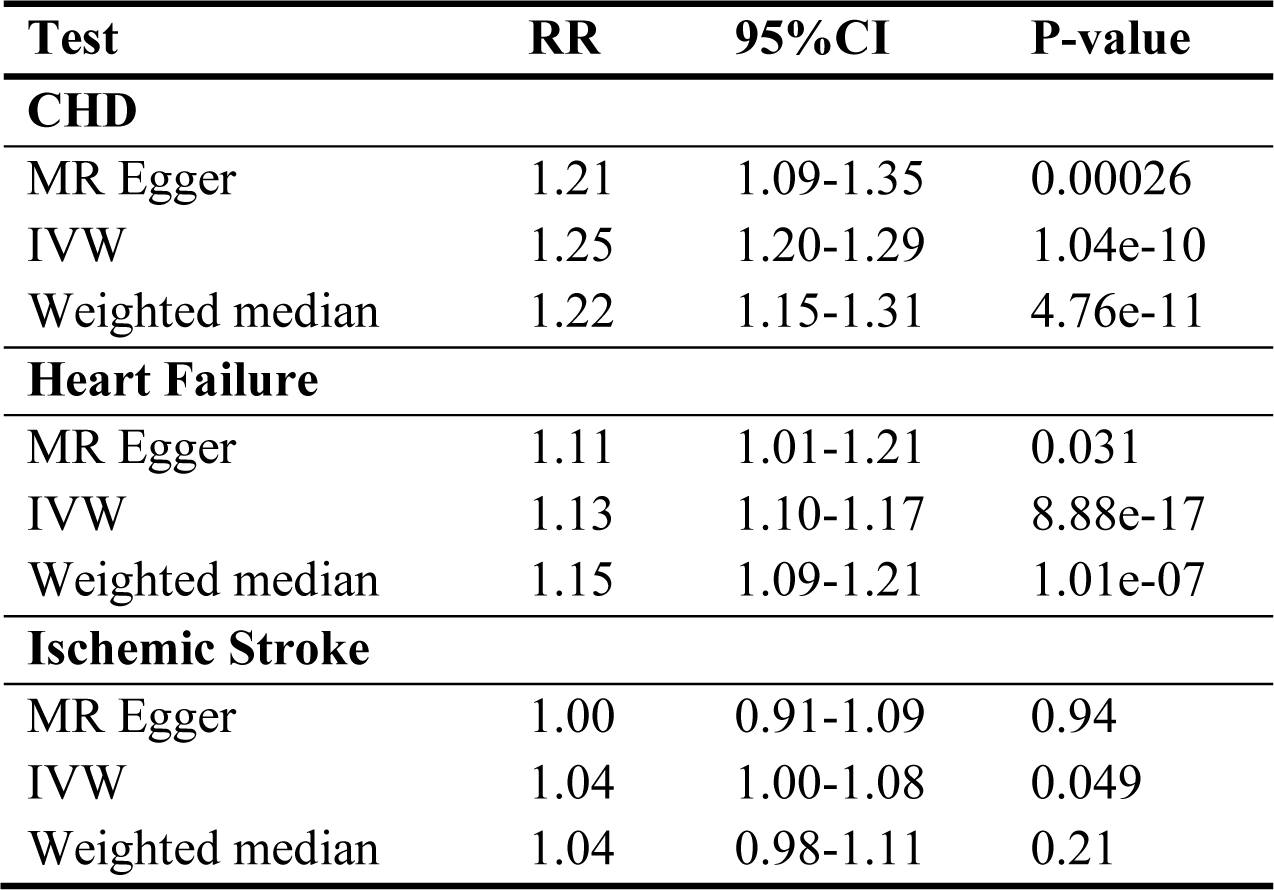
Mendelian randomization (MR) study on the causal link between the metabolic syndrome (MetS) and the three major CVD; coronary heart disease (CHD), heart failure, and ischemic stroke. IVW=Inverse-variance weighted.

| **Test** | **RR** | **95%CI** | **P-value** |
| --- | --- | --- | --- |
| **CHD** |  |  |  |
| MR Egger | 1.21 | 1.09-1.35 | 0.00026 |
| IVW | 1.25 | 1.20-1.29 | 1.04e-10 |
| Weighted median | 1.22 | 1.15-1.31 | 4.76e-11 |
| **Heart Failure** |  |  |  |
| MR Egger | 1.11 | 1.01-1.21 | 0.031 |
| IVW | 1.13 | 1.10-1.17 | 8.88e-17 |
| Weighted median | 1.15 | 1.09-1.21 | 1.01e-07 |
| **Ischemic Stroke** |  |  |  |
| MR Egger | 1.00 | 0.91-1.09 | 0.94 |
| IVW | 1.04 | 1.00-1.08 | 0.049 |
| Weighted median | 1.04 | 0.98-1.11 | 0.21 |

### Polygenic risk scores

As could be seen in figure 4, the PRS for CHD was increased in all groups of combinations of MetS components, except for the group with the combination of the GLU and TG criteria. Generally, the PRS for CHD increased with increasing number of MetS criteria, but the group with all 5 criteria did not show higher PRS for CHD when compared with three of the groups with four components (not including TG or BP or WC), while it was higher compared with two of the groups with four components (not including GLU or HDL). The group with the combination of GLU+WC+HDL showed the same estimate for the PRS for CHD as the group with all 5 components.

**Figure 4.**
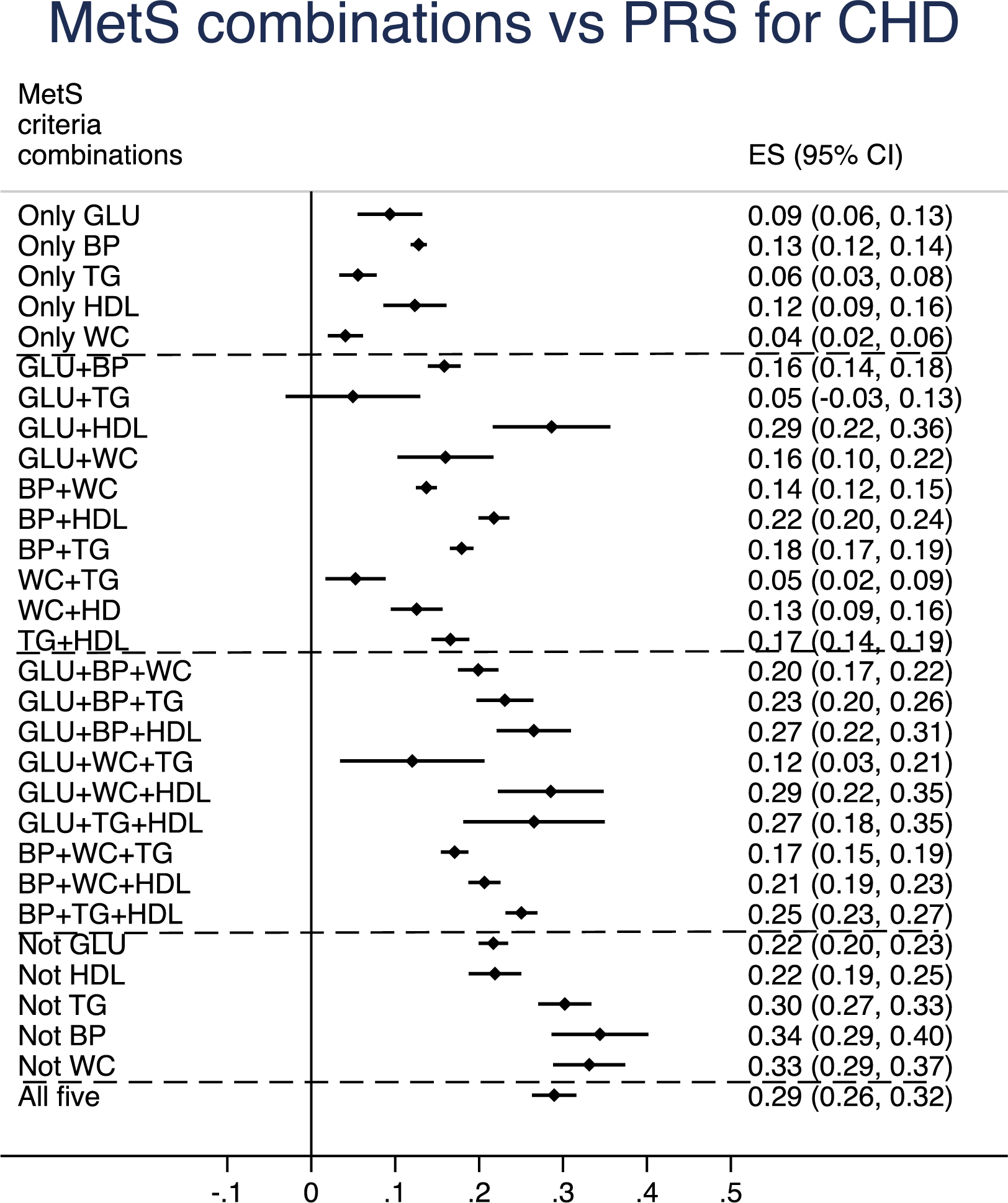
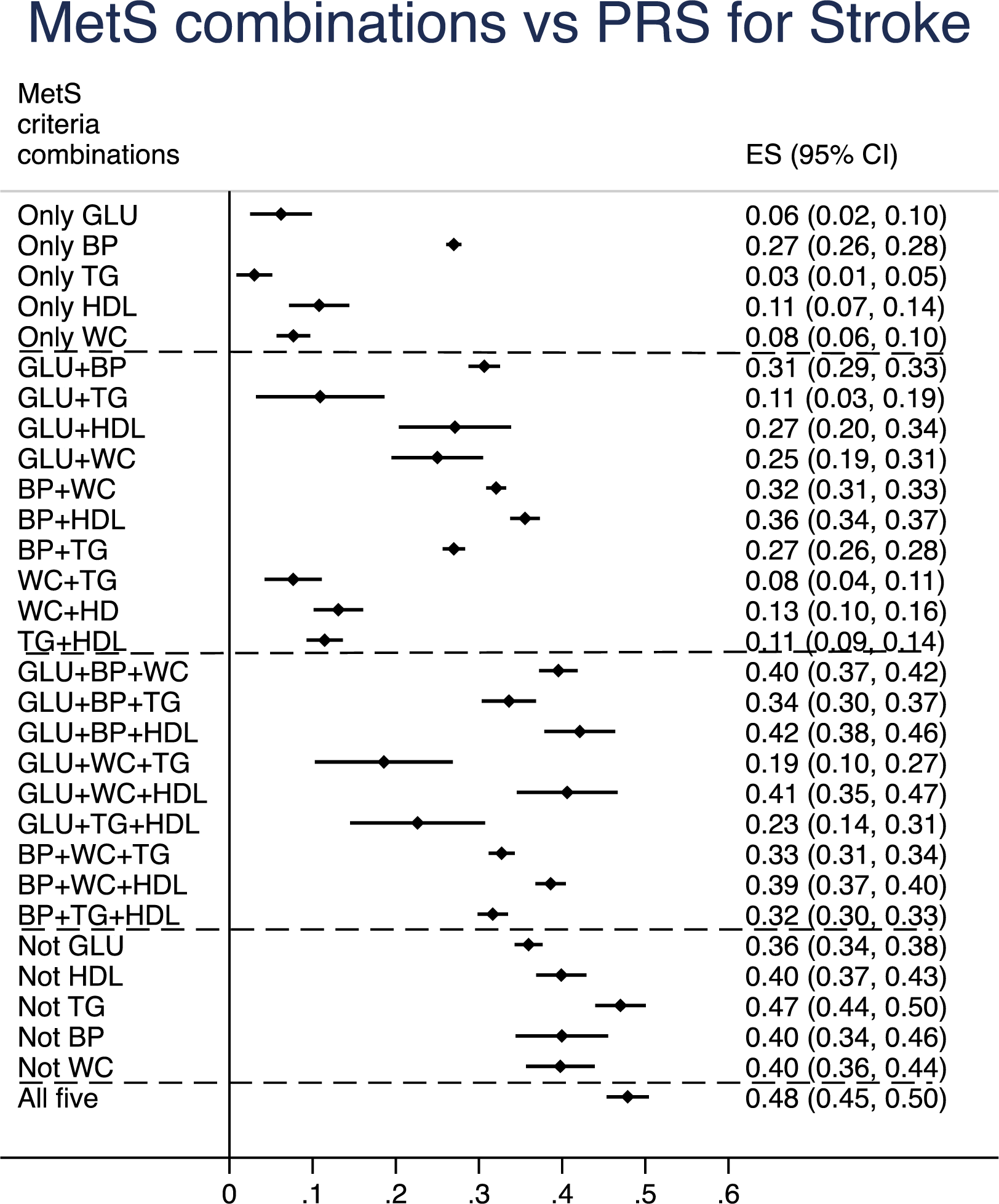
Relationships between different combinations of metabolic syndrome (MetS) components and a polygenetic risk score (PRS) for coronary heart disease (CHD, upper panel) and a PRS for ischemic stroke (lower panel). Betas and 95%CI are given. The group without any MetS components was use as the reference group.

A rather similar pattern was seen groups of combinations of MetS components were related to the PRS for ischemic stroke. All groups showed an increased RPS compared to the groups without any MetS components. For this outcome, the group with all 5 components showed a higher PRS for stroke than three of the groups with four components (not including GLU or HDL or WC) and a higher estimated (although not significant) compared to the group with the combination of GLU+WC+HDL.

## DISCUSSION

The present study first confirmed previous knowledge on an increased risk of future CVD in individuals with MetS, and an increasing risk with increasing number of MetS criteria. Novel findings were derived from the fact that the UK biobank both provide a large sample size and a sufficiently long follow-up period to collect incident CVD data, which permits division into MetS criteria combinations with a good power. It was then found that certain MetS criteria combinations were associated with a higher risk than others, and that some combinations with three or four criteria showed a similar increased risk as seen in individuals with all five criteria. We could also show with both observational data and Mendelian randomization that MetS was linked to MI and HF, but less so to ischemic stroke. Using derived PRS it could be shown that almost all combinations of MetS criteria were related to an increased genetic risk for CHD and ischemic stroke, with a pattern being similar to the observational data.

It has been shown multiple times that MetS, and the number of MetS components, is related to future CVD and/or mortality ^3, 5–17^, so this part of the study was mainly used to validate that the sample of the UK biobank behaved in the expected way, and that the use of non-fasting values for glucose and triglycerides (with some slight corrections for fasting time) were appropriate to use.

The primary aim of this study was however to evaluate if certain combinations of MetS criteria were associated with a higher risk of CVD than others. Figure 3 clearly shows this was the case. Within the groups with combinations of three MetS criteria, the GLU+TG+HDL criteria group showed the highest risk with the lowest 95%CI limit not overlapping with five of the other groups with combinations of three MetS criteria. Since the HR for this group is as high as the HR seen in the group with all five criteria, this particular combination seems to also capture the risk associated with abdominal obesity and a high blood pressure. This group also showed pronounced relationships vs the PRS for CHD and ischemic stroke suggesting causal relationships. Thus, the present study confirms the previous critique that MetS does not take into account different combinations of the criteria, since it is obvious that some combinations of MetS criteria were associated with a higher risk of CVD than others.

An interesting finding in the present study was that the patterns of risk estimates for the different combinations of MetS criteria were rather similar for MI and HF, but for ischemic stroke the associations were generally less strong and other patterns were seen. It is well known from observational studies that blood pressure is by far the most powerful risk factor for stroke, while many risk factors contribute to the risk of myocardial infarction and heart failure. A recent multi-variate MR study with multiple risk factors showed a similar view ^24^. Thus, it cannot be expected that MetS would be as strongly related to ischemic stroke as compared to the other two investigated CVDs. This assumption is confirmed in the present study, in which MR was used to evaluate a possible causal role of MetS regarding the three CVDs of interest.

The major strength of the present study is the large sample size and great number of incident cases of CVDs that made it possible to perform the division in the many groups with combinations of MetS components with still a decent power.

For the MR part we did use a GWAS of MetS conducted in the UK biobank ^23^ and therefore we did not use the most recent GWAS studies of the three CVDs including a large number of UK biobank cases in order to avoid overfitting problems. In the HERMES consortium of heart failure, about 15% of the cases were derived from UK biobank, but this would likely not induce any major overestimation of the causal relationship.

In conclusion, certain combinations of risk factors in individuals with the metabolic syndrome criteria showed a higher risk of future CVD than others. A combination of high glucose, low HDL, and high triglycerides seems to capture the highest risk of CVD.

## SUPPLEMENTAL MATERIAL

Suppl Table 1

## Data Availability

All data produced in the present study are available upon reasonable request to the authors

## ABBREVIATIONS

BP: Blood pressure
CHD: Coronary heart disease
CVD: Cardiovascular disease
GLU: Glucose
GWAS: Genome-wide association studies
HF: Heart failure
IVW: Inverse-variance weighted meta-analysis
MetS: Metabolic syndrome
MI: Myocardial infarction
MR: Mendelian randomization
TG: Triglycerides
WC: Waist circumference

